# Differences in maternally inherited and age-related de novo mitochondrial DNA variants between ART and spontaneously conceived individuals associate with low birth weight

**DOI:** 10.1101/2022.09.05.22279608

**Authors:** Joke Mertens, Florence Belva, Aafke P.A. van Montfoort, Filippo Zambelli, Sara Seneca, Edouard Couvreu de Deckersberg, Maryse Bonduelle, Herman Tournaye, Katrien Stouffs, Kurt Barbé, Hubert Smeets, Hilde Van de Velde, Karen Sermon, Christophe Blockeel, Claudia Spits

## Abstract

**Background:** Children born using assisted reproductive technologies (ART) have an increased risk of a lower birth weight, the cause of which remains unclear. As a causative factor, we hypothesized that variants in the mitochondrial DNA (mtDNA) that are not associated with disease, may explain changes in birth weight.

**Methods:** We deep-sequenced the mtDNA of 451 ART and spontaneously conceived (SC) individuals, 157 mother-child pairs and 113 individual oocytes from either natural menstrual cycles or cycles with ovarian stimulation (OS). The mtDNA genotypes were compared across groups and logistic regression and discriminant analysis were used to study the impact of the different factors on birth weight percentile.

**Results:** ART individuals more frequently carried variants with higher heteroplasmic loads in protein and rRNA-coding regions. These differences in the mitochondrial genome were also predictive of the risk of a lower birth weight percentile, irrespective of the mode of conception but with a sex-dependent culture medium effect. The higher incidence of these variants in ART individuals results both from maternal transmission and *de novo* mutagenesis, which we found not to be caused by OS but to be associated to maternal ageing.

**Conclusions:** MtDNA variants in protein and rRNA coding regions are associated with a lower birth weight and are more frequently observed in ART children. We propose that these non-disease associated variants can result in a suboptimal mitochondrial function that impacts birth weight. Future research will establish the long-term health consequences of these changes and how these findings will impact the clinical practice and patient counselling in the future.

## INTRODUCTION

The birth of Louise Brown in 1978 heralded the start of a revolution in reproductive medicine that has made parenthood possible for couples previously considered untreatably infertile. Since then, assisted reproductive technologies (ART) have gone through considerable developments and its use has increased exponentially, with over 8 million children born worldwide. Over the past decades, virtually all steps of the process have undergone continuous improvement. Nevertheless, concerns about the safety of these procedures for the children born after ART have always been present, and the systematic study of large cohorts of children over many years has revealed that these individuals have indeed an adverse outcome as compared to those born after spontaneous conception (SC). There is by now much evidence that ART children are at risk of being born small for gestational age^1^ and there is an increasing number of studies showing that these children are at risk for an abnormal cardio-metabolic profile later in life^2^.

Over the years a large body of research has been devoted to the identification of the causes and molecular basis of these adverse effects in ART children, based on the knowledge that suboptimal early development conditions have a significant impact on the future health of the individual^3^. In this context, the vast majority of studies have focused on the search for ART-induced epigenetic changes, both in individuals and in placental samples. Although a significant number of animal studies have demonstrated a link between epigenetic alterations and different aspects of ART, studies in human did not provide conclusive results^4^. Also, a recent study showed that *in vitro* fertilization does not increase the incidence of chromosomal abnormalities in human fetal and placental lineages, and did not find a link between these abnormalities and birth weight^5^.

In this study, we hypothesized these differences to be caused by mitochondrial DNA (mtDNA) variants. In the general population, low birth weight is associated with metabolic abnormalities^6^, which in adulthood have been linked to mitochondrial dysfunction^7^. Furthermore, inherited mtDNA defects can predispose to obesity and insulin resistance^8^ and low birth weight is common in individuals born with mitochondrial DNA disease^9,10^. Female subfertility and ovarian stimulation (OS) strongly associate with the risk for low birth weight in ART children^11,12^, and may both be linked to mitochondrial function as oxidative phosphorylation plays a crucial role during gametogenesis as well as in female subfertility^13^.

We hypothesized that subfertile women more frequently carry mtDNA variants which negatively affect mitochondrial function and contribute to their subfertility. Furthermore, work in animal models suggests that OS increases the frequency of mtDNA deletions in the oocytes^14,15^ and affects mitochondrial function^16^. Therefore, we postulated that maternal transmission of subfertility-associated mtDNA genotypes and *de novo* mutagenesis during OS could result in adverse perinatal outcomes in children carrying these mtDNA genotypes.

To test these hypotheses, we studied the mtDNA of 270 ART and 181 spontaneously conceived (SC) children, 157 mother-child pairs and 113 oocytes donated in both natural menstrual cycles and after OS by the same donors and investigated the link between mtDNA variants and birth weight.

## METHODS

### Recruitment and donor material

Table 1 lists the details on children and mothers included in this study. The participants were recruited at the Universitair Ziekenhuis Brussel (UZ Brussel, Belgium) and at the Maastricht University Medical Centre (Netherlands). For the study of oocytes, twenty-nine young fertile women donated oocytes in up to three natural menstrual cycles and after one OS cycle. All donors were recruited at the Center for Reproductive Medicine of the UZ Brussel, where all medical procedures took place. The inclusion criteria for the recruitment were: 19-35 years old, healthy, preferably with a proven fertility, body mass index of 18-32. The exclusion criteria were: any type of medical or genetic condition that may interfere with the health of the oocytes, endometriosis grade AFS III-IV, ovulatory disorders belonging to WHO categories 1-6, body mass index of <18 or >32, use of medication that may interfere with the oocyte competence and psychiatric disorders. The donors underwent two types of donation cycles: up to three natural cycles and one cycle after ovarian stimulation (OS, Supplementary Table S1). In the natural cycle, a blood sample was taken on day 2 of the follicular phase to assess several parameters (serum estradiol, progesterone, luteinizing hormone, follicle-stimulating hormone (FSH), and human chorionic gonadotrophin (hCG) levels). On day 10, a blood sample was taken again to check the same parameters except hCG as well as a vaginal ultrasound to assess follicular development. If not sufficient, additional blood samples and ultrasound were taken. The oocyte retrieval was performed 32 hours after administration of 5000 IU exogenous hCG. The ovarian stimulation cycle started on the first day of menstruation following the natural cycle. Starting on day 2 of the cycle, a gonadotrophin-releasing hormone (GnRH) antagonist protocol was applied, using recombinant FSH if hormonal analysis were within normal limits on day 2. The starting dose of the recombinant FSH varied between 150-300 IU per day. At day 6 of stimulation, a GnRH antagonist (Ganirelix; Orgalutran®; MSD, Oss, The Netherlands) with a dose of 0.25 mg per day was given. Final oocyte maturation was achieved by administration of a GnRH agonist (Gonapeptyol®, Ferring Pharmaceuticals, St-Prex, Switzerland) with a dose of 0.2 ml when at least 3 follicles reached a mean diameter of 17-20 mm. Oocyte retrieval was performed according to routine procedures, 36 hours after trigger.

**Table 1.** Characteristics of the ART and SC individuals included in this study.

|  | ART<br>(N=270)<br>% (N) | SC<br>(N=181)<br>% (N) | P-value |
| --- | --- | --- | --- |
| **Source material** |  |  |  |
| Blood<sup>1</sup> | 49.8% (103) | 35.9% (65) |  |
| New-borns | 17.0% (46) | 0.0% (0) |  |
| Young adults | 21.1 (57) | 35.9% (65) |  |
| Placenta<sup>2</sup> | 31.9% (66) | 28.2% (51) |  |
| Buccal cells<sup>3</sup> & Saliva<sup>2</sup> | 37.4% (101) | 35.9% (65) |  |
| Children | 37.4% (101) | 2.8% (5) |  |
| Young adults | 0.0% (0) | 33.1% (60) |  |
| Mother-child pairs | 67 | 115 |  |
| **Clinical parameters** |  |  |  |
| Mean maternal age<sup>4</sup> ± SD* | 32.5 ± 3.7 (211) | 30.1 ± 4.4 (173) | <0.0001<sup>a</sup> |
| Maternal BMI >25* | 40.2% (86/214) | 25.6% (42/164) | 0.0031<sup>b</sup> |
| Birth weight* | 224 | 164 |  |
| Mean (grams) ± SD | 3377.9 ± 536.0 | 3394.3 ± 500.7 | 0.7596<sup>a</sup> |
| <10th percentile | 7.0% (19) | 5.0% (9) | 0.2407<sup>b</sup> |
| <25th percentile | 15.2% (41) | 14.4% (26) | 0.4992<sup>b</sup> |
| Preterm birth (<37 weeks)* | 5.8% (13/224) | 6.1% (10/164) | 1.0000<sup>b</sup> |
| Sex* | 41.5% (112) – | 44.8% (81) – | 0.3176<sup>b</sup> |
| (Male – Female) | 41.5% (112) | 55.2% (100) |  |
| Culture medium | 224 | NA |  |
| Vitrolife G3 | 19.3% (52) | NA |  |
| Vitrolife G5 | 24.4% (66) | NA |  |
| Cook | 18.1% (49) | NA |  |
| UZB | 21.1% (57) | NA |  |
| Unknown | 17.0% (46) | NA |  |
| Gestational hypertension* | 4.1% (8/193) | 1.9% (3/159) | 0.3571<sup>b</sup> |
| Gestational diabetes* | 1.0% (2/193) | 1.3% (2/158) | 1.0000<sup>b</sup> |
| Primiparity* | 75.2% (167/222) | 47.2% (68/144) | <0.0001<sup>b</sup> |
| Smoking during pregnancy* | 5.5% (12/220) | 7.7% (13/169) | 0.4086<sup>b</sup> |

The donors were recruited in different centers: <sup>1</sup>UZ Brussel, Belgium, <sup>2</sup>Maastricht University Medical Center, The Netherlands and <sup>3</sup>Faculty of Medicine and Pharmacy, Vrije Universiteit Brussel, Belgium. <sup>4</sup>Age in years. \*Only a subset of samples was provided with clinical data such as maternal age, maternal BMI (body mass index), birth weight (categorized in percentiles), gestational age, sex, culture medium, complications during pregnancy and parity. UZB medium is an in-house medium made in the UZ Brussel. All individuals are singletons from fresh embryo transfers. Some characteristics were not applicable (NA) for some samples. N indicates number of samples for which this data was available. Statistical tests were performed using the student t test (<sup>a</sup>) or the Fisher's exact test (<sup>b</sup>).

All studies have been approved by the local ethical committee of the UZ Brussel and the ethical review board of the Maastricht University Medical Centre. All individuals signed an informed consent.

### DNA isolation and long-range PCR protocol for mtDNA enrichment

DNA was extracted from peripheral blood, placenta, saliva and buccal swab samples, and isolated from single oocytes as previously described^17,18^. The mtDNA was enriched by long-range PCR using two primer sets to cover the full mtDNA in bulk DNA samples. For single oocytes, only one primer set was used. The sequences of the first primer set (5042f-1424r) were 5′-AGC AGT TCT ACC GTA CAA CC-3′ (forward) and 5′-ATC CAC CTT CGA CCC TTA AG-3′ (reverse) generating amplicons of 12.9 Kb. The sequences of the second primer set (528f-5789r) were 5′-TGC TAA CCC CAT ACC CCG AAC C-3′ (forward) and 5′-AAG AAG CAG CTT CAA ACC TGC C-3′ (reverse) generating amplicons of 5.3 Kb. Both primer sets were purchased from Integrated DNA technologies and were diluted to 10 µmol and kept at −20°C. The master mix for the long-range PCR was prepared as follows: 10 µL of 5x LongAmp buffer (LongAmp Taq DNA Polymerase kit M0323L, New England Biolabs, stored at −20°C), 7.5 µL dNTPs (dNTP set, IllustraTM, diluted to 2 mM and stored at −20°C), 2 µL of the forward and reverse primers (10 µM), 2 µL of Taq Polymerase LongAmp (5 units) and 50 ng of diluted DNA (working solution: 10 ng/µL) in a total of 50 µL. In case of the oocytes, a single-cell long-range PCR was applied. Here, only the first primer set was used and 2.5 µL of Tricine (Sigma-Aldrich T9784, diluted to 200 nM and stored at 4°C) was added to buffer the alkaline lysis buffer (200 mM NaOH and 50 mM DTT) used in the oocyte collection. The master mix was directly added to the sample. Successful PCR amplification was confirmed by running a gel-electrophoresis. This method was thoroughly validated with a lower detection threshold with 2% for single cells and 1.5% for bulk material. Specificity of the primer sets for the mitochondrial genome was controlled by amplifying DNA of RhoZero cells, which do not contain mitochondrial DNA. More detailed protocols of the long-range PCR are published previously^17,19^.

### Preparation of samples for sequencing and data analysis

After PCR amplification, the samples were purified with AMPure beads and the amplicons from both primer sets were pooled together in a 0.35/0.65 ratio (35% of the amplicon from the second primer set and 65% of the amplicon of the first primer set). Next, the library was prepared with the KAPA HyperPlus kit and purified with AMPure beads. The quality of the library was checked by electrophoresis on the AATI Fragment Analyzer using the HS NGS Fragment kit. Next, the libraries were diluted to 2 nM in EBT buffer and pooled before loading on the Illumina platform. Alignment to the reference genome (NC_0.12920.1) was done using BWA-MEM. The generated bam files were uploaded to mtDNA server^20^ and MuTect^21^ was used to detect small insertions and deletions. Variants with >1.5% heteroplasmic load, or >2% in case of the single oocytes, were annotated and possible amino-acid changes were identified using MitImpact2^22^. Homoplasmies were defined as variants with a load >98.5% (100% minus the detection limit) for the samples of the children and the mothers, for the oocytes this was >98%. Regions of the mtDNA known to be prone to PCR and sequencing error were excluded (Supplementary Table S2). The variants identified in all the samples included in this study are listed in the supplementary data file.

### Statistics

Statistical analysis was carried out using SPSS (IBM). The heteroplasmic variants were analysed through an orthogonally rotated exploratory factor analysis, which extracts the variability among variables in the form of independent latent variables (more details can be found in the supplementary methods). Binary logistic regression and discriminant analysis were used to build models to predict the effect of different variables on the birth weight percentile (more details on the models can be found in the supplementary methods). For these, samples with missing data were excluded from the analyses. Other statistical analyses were done using the Fisher’s exact, Mann-Whitney U and Student t-test and p-values <0.05 were considered significant. Correcting for multiple testing was done using the Bonferroni method, and the adjusted threshold for significance is indicated where relevant.

## RESULTS

### Individuals born after ART display a different mitochondrial DNA variant genotype than their spontaneously conceived peers

First, the data was controlled for differences in haplogroup distribution. No significant differences were found in haplogroup and subhaplogroup distribution across ART and SC individuals (Figure 1a, and Table S3 and S4 in supplementary appendix). Next to the haplogroup variants, most individuals carried additional homoplasmic variants, which were categorized according to their location in the mitochondrial genome and their impact on the amino acid sequence: hypervariable region (HV), non-coding, origin of replication on the heavy strand (OHR), termination-associated sequence (TAS), rRNA, tRNA, synonymous and non-synonymous protein-coding variants. The proportion of individuals carrying homoplasmic variants was similar in both groups, as well as the location of the variants (Figure 1b and Table S5 in supplementary appendix).

**Figure 1.**
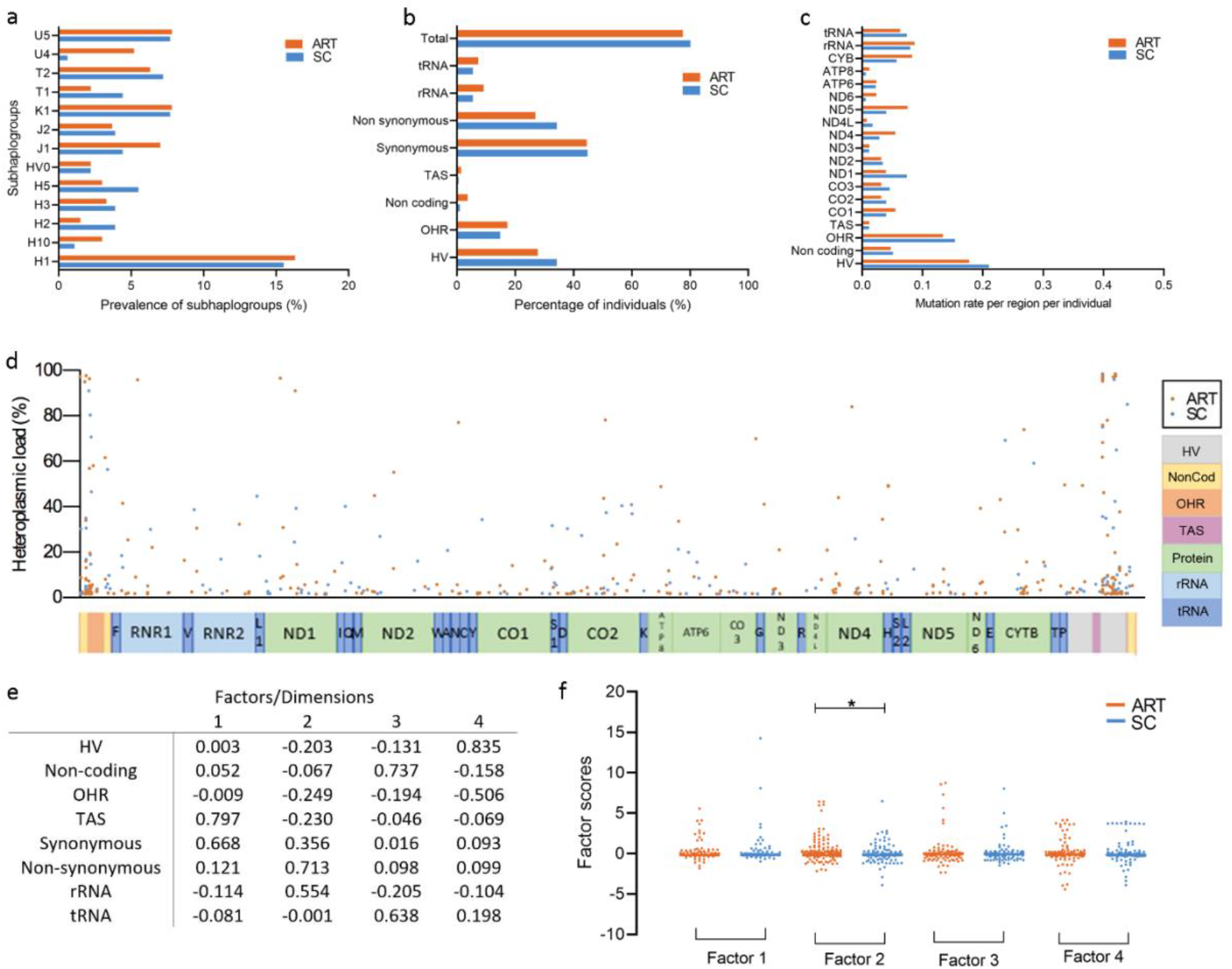
Individuals born after ART carry a different mitochondrial DNA variant landscape compared to their spontaneously conceived peers. **a**. Prevalence of the subhaplogroups across ART and SC individuals. Subhaplogroup U4 appeared more frequent in the ART than in the SC group, but after correcting for multiple testing, this was not statistically significant (5.2% vs 0.6%, Fisher’s exact test with Bonferroni correction, p=0.007 (p values ≤ 0.0038 were considered significant)). **b**. Percentage of ART and SC individuals with homoplasmic variants in different categories. Percentages are not adding up to 100% because one individual can have multiple variants. **c**. Mutation rate per region, per individual in ART and SC individuals, calculated as the number of variants identified in the region divided by the total number of individuals per group. **d**. Overview of all heteroplasmic variants identified, plotted on the mitochondrial genome according to their heteroplasmic load. **e**. Component matrix generated by the factor analysis using the sum of the heteroplasmic loads per individual in eight different categories. **f**. Factor scores of the heteroplasmic data of ART and SC individuals. ART individuals more frequently carry higher scores for factor 2 (*Mann-Whitney, p=0.021). ART: assisted reproductive technologies, SC: spontaneously conceived, HV: hypervariable region, OHR: origin of replication on the heavy strand, TAS: termination associated sequence. Synonymous and non-synonymous variants are subcategories of protein-coding variants.

In total, 430 heteroplasmic variants were identified, with on average 0.94 and 0.97 variants per individual in the ART and SC samples, respectively. There were no differences in the distribution of the variants according to their location and type of nucleotide substitution (Figure 1c, and Table S6 and S7 in supplementary appendix). An overview of all identified heteroplasmic variants is shown in Figure 1d. 66.0% of these variants were unique to one individual and individuals frequently carried multiple variants with different heteroplasmic loads. To factor in the heteroplasmic load and to reduce the data complexity due to the diverse location of the variants, we added up the loads of the heteroplasmic variants per individual, categorized per location (HV, tRNA, etc). This resulted in a matrix of “sums of heteroplasmic loads” per each individual, per region. ART and SC individuals did not differ in the mean sum of heteroplasmic loads per location in their mtDNA (Table S8 in supplementary appendix). Next, we used an orthogonally rotated exploratory factor analysis on the sum of the heteroplasmic loads of each location in the mtDNA per individual. This further reduced the complexity of the data to four factors; the component matrix can be found in Figure 1e. These factors reflect covariance between types of mtDNA variants. We extracted the scores for each factor for all samples and used these to further explore diversity in heteroplasmic variant composition across the two groups. We controlled for the use of different tissues, the age at sampling and the maternal age at conception, and found no significant differences in the scores of any of the four factors across the different tissues (Figure S1a in supplementary appendix), between the ages at sampling (Figure S1b in supplementary appendix) nor a correlation with maternal age at conception (Figure S1c in supplementary appendix). Conversely, ART individuals were found to have significantly higher scores in factor 2, driven by the presence of protein-coding and rRNA variants and lack of HV, OHR and TAS variants, as compared to SC children (Mann-Whitney test, p=0.021). Factors 1, 3 and 4 were not different between the two groups (Figure 1f). This indicates that individuals born after ART tend to carry higher loads of heteroplasmic variants in the protein-coding and/or rRNA regions, in combination with lower or similar heteroplasmic loads in the other regions compared to their SC peers.

### Differences in the mtDNA variant profile correlate with birth weight percentiles

We investigated the association between the birth weight of 388 individuals and their mtDNA profile (164 SC and 224 ART individuals). The birth weight was adjusted for gestational age and sex and categorized as under or above the 10^th^ percentile, where <P10 is considered small for gestational age, and birth weight under or above the 25^th^ percentile.

There was no association between haplogroup and birth weight percentile (Figure 2a and Table S9 in supplementary appendix) nor with the presence of homoplasmic variants (Figure 2b, Table S10 in supplementary appendix) between individuals <P10 or >P10, regardless of mode of conception. Conversely, SC individuals who were <P10 or <P25 had significantly higher scores in factor 2 of the heteroplasmic variants analysis than children born at >P10 or >P25 (Man-Whitney test, <P10; p=0.04; <P25: p=0.003, Figure 2c and 2d). In line with this, SC <P10 and <P25 individuals more frequently carried variants in protein and rRNA loci (<P10: 77.8% and >P10: 34.8%, Fisher’s exact test, p=0.0138, odds ratio: 6.55, 95% confidence interval (CI) 1.31-32.61, and <P25: 65.4% and >P25: 31.9%, Fisher’s exact test, p=0.0018, odds ratio 4.04, 95% CI 1.67-9.77). Strikingly, this difference was not observed in the ART group as a whole (Fisher’s exact test, p=1 for both percentiles of birth weight). Stratification by sex revealed that only <P25 ART females followed a similar pattern as SC individuals, although not statistically significantly (Fisher’s exact test, p=0.2075, Figure 2e-f).

**Figure 2.**
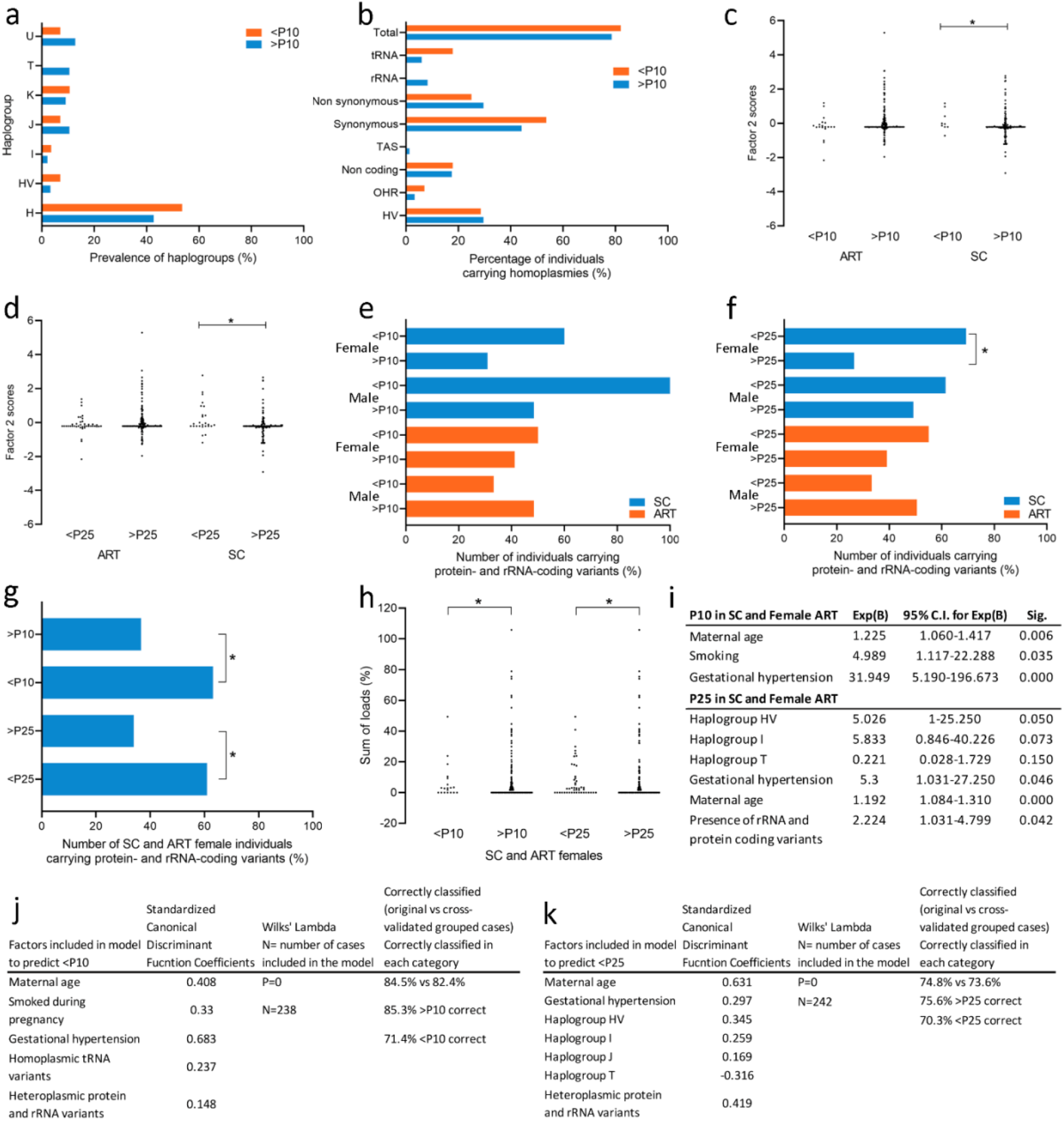
Differences in mtDNA variant composition correlate to birthweight. **a**. Prevalence of the haplogroups across ART and SC individuals with a birth weight percentile <P10. **b**. Percentage of individuals carrying homoplasmic variants in the different regions and categorized according to whether they were under or above P10. **c**. Factor 2 scores in ART and SC children categorized according to whether they were under or above P10. SC children with a birth weight <P10 had significantly higher scores for factor 2 (*Mann-Whitney test, p=0.04). **d**. Factor 2 scores in ART and SC children categorized according to whether they were under or above P25. SC children with a birth weight <P25 had significantly higher scores for factor 2 (*Mann-Whitney test, p=0.003). **e**. Number of ART and SC individuals, stratified for sex and birth weight percentile under or above P10, carrying variants in the protein and rRNA loci (Total number of samples <P10: ART females: N=10/112, ART males: N=9/112, SC females: N=5/92 and SC males: N=4/72). **f**. Number of ART and SC individuals, stratified for sex and birth weight percentile under or above P25, carrying variants in the protein and rRNA loci. Female SC children <P25 more frequently carry protein and rRNA variants than SC children >P25 (Fisher’s exact test, p=0.004; Total number of samples <P10: ART females: N=20/112, ART males: N=21/112, SC females: N=13/92 and SC males: N=13/72). **g**. SC and female ART individuals, stratified for birth weight under or above P10 and under or above P25, carrying variants in the protein and rRNA loci. Both SC and female ART individuals <P10 and <P25 more frequently carried protein and rRNA variants than female SC individuals >P10 or >P25 (Fisher’s exact test, P10: p=0.028, P25: p=0.0008). **h**. Sum of loads of SC and female ART children under or above P10 and P25 carrying variants in the protein- and rRNA-coding regions. Both <P10 and <P25 SC and female ART children carried higher sum of loads of protein- and rRNA coding variants (Fisher’s exact test, P10: p=0.036 and P25: p=0.0009) **i**. Result of the binary logistic regression for the P10 and P25 SC and female ART individuals. In both models, maternal age, smoking and gestational hypertension had the most influence on the outcome of birth weight percentile. Additionally, the model for P25 also included the presence of haplogroups HV and I, the absence of haplogroup T and the presence of protein and rRNA variants. **j-k**. Result of the discriminant analysis for the P10 (j) and P25 (k) SC and female ART individuals. In both models maternal age, gestation hypertension and the presence of heteroplasmic protein and rRNA variants had the most influence on the outcome of birth weight percentile. In the P10 model, smoking during pregnancy and the presence of homoplasmic tRNA variants were also included. In the P25 model, haplogroups HV, I, J and T were added. ART: assisted reproductive technologies, SC: spontaneously conceived, HV: hypervariable region, OHR: origin of replication on the heavy strand, TAS: termination associated sequence. Synonymous and non-synonymous variants are subcategories of protein-coding variants.

A potential explanation to this remarkable difference between male and female ART individuals can be sex-related differences in sensitivity to different embryo culture mediums. Our study cohort partially overlaps with that of a study showing that embryos that have been grown in Cook medium have lower birth weights, and that male embryos are more sensitive to this effect^22^. We observe a similar trend in the present study, where particularly male individuals that were subjected to Cook and UZB medium (e.i. an in-house medium made in the UZ Brussel) are more frequently born with a birth weight <P25 than females (25.5% in males vs 10.7 % in females). Remarkably, only 7.7% of these <P25 males carried variants in protein- and rRNA-coding loci as compared to 30.0% of the <P25 females. Regrettably, the small size of this subgroup of samples does not allow for a conclusive and statistically significant analysis of the interactions between culture medium, sex, mtDNA variants and birth weight. Therefore, in further analysis related to birth weight, we excluded male ART individuals to avoid the bias caused by culture media. With this in mind, we found that SC and female ART individuals that were <P10 or <P25 more frequently carried protein and rRNA variants than when they were born >P10 and >P25 (<P10: 63.2% and >P10: 36.6%, Fisher’s exact test, p=0.028, odds ratio 2.97, 95% CI 1.13-7.81, and <P25: 60.9% and >P25: 33.9%, Fisher’s exact test, p=0.0008, odds ratio 3.03, 95% CI 1.58-5.82, Figure 2g). Additionally, SC and female ART individuals with a birth weight <P10 and <P25 had overall higher cumulative heteroplasmic loads of protein and rRNA variants (Mann-Whitney test, p=0.036 and p=0.0009 respectively, Figure 2h).

Next, we used a backward stepwise conditional binary logistic regression to study the effect of the different mtDNA-related variables on the birth weight, including the potential confounding factors listed in table 1. We included in the model any parameter that showed through univariate analysis an association to birth weight percentile with p<0.2 (Table S11 in supplementary appendix). For <P10, only maternal age, smoking during pregnancy and gestational hypertension showed a statistically significant impact on birth weight (Figure 2i). In the case of <P25, the logistic regression resulted in a model including three haplogroups and gestational hypertension, maternal age and the presence of heteroplasmic protein- and rRNA-coding variants. The three last had all a significant impact in increasing the chances of being born <P25 (Figure 2i). Finally, discriminant analysis using the same variables resulted in two statistically significant models (Wilk’s Lambda p=0) that were able to correctly classify <P10 or >P10 individuals in 84.5% of cases, and <P25 or >P25 in 74.8% of cases (Figure 2j-k).

### The differences between ART and SC individuals are due to both maternally transmitted and *de novo* variants

To investigate the origin of these differences in the mtDNA, we studied 67 ART and 90 SC mother-child pairs. We categorized a variant as “transmitted” when it was present in the mother and in the child, and “*de novo*” when the variant was only present in the child.

We found a higher incidence of ART mother-child pairs presenting *de novo* variants in the protein-coding regions than SC mother-child pairs (ART: 38.8% (26/67), SC: 20.0% (18/90), Fisher’s exact test, p=0.01; odds ratio: 2.54, 95% CI 1.24-5.17, Figure 3a). We generated the sum of the heteroplasmic loads of the transmitted and *de novo* variants per region of each of the children and submitted them to factor analysis using the matrix in Figure 1e. The factor scores of the mothers correlated highly with those of their child (Figure S2 in supplementary appendix). The scores for factor 2 (driven by the presence of protein-coding and rRNA variants) were significantly higher in ART individuals for the transmitted but not for the *de novo* variants (Mann-Whitney test, transmitted: p=0.04, *de novo*: p=0.08; Figure 3b and Figure 3c respectively). Furthermore, the scores for factor 1 (synonymous and TAS variants) were significantly higher for the *de novo* variants in the ART individuals (Mann-Whitney test, p=0.03, Figure 3c). These findings suggest that the differences in mtDNA variants between ART and SC children are due to a higher transmission of protein- and rRNA-coding variants by ART mothers, in combination with a more frequent acquisition of *de novo* protein-coding variants in ART children.

**Figure 3.**
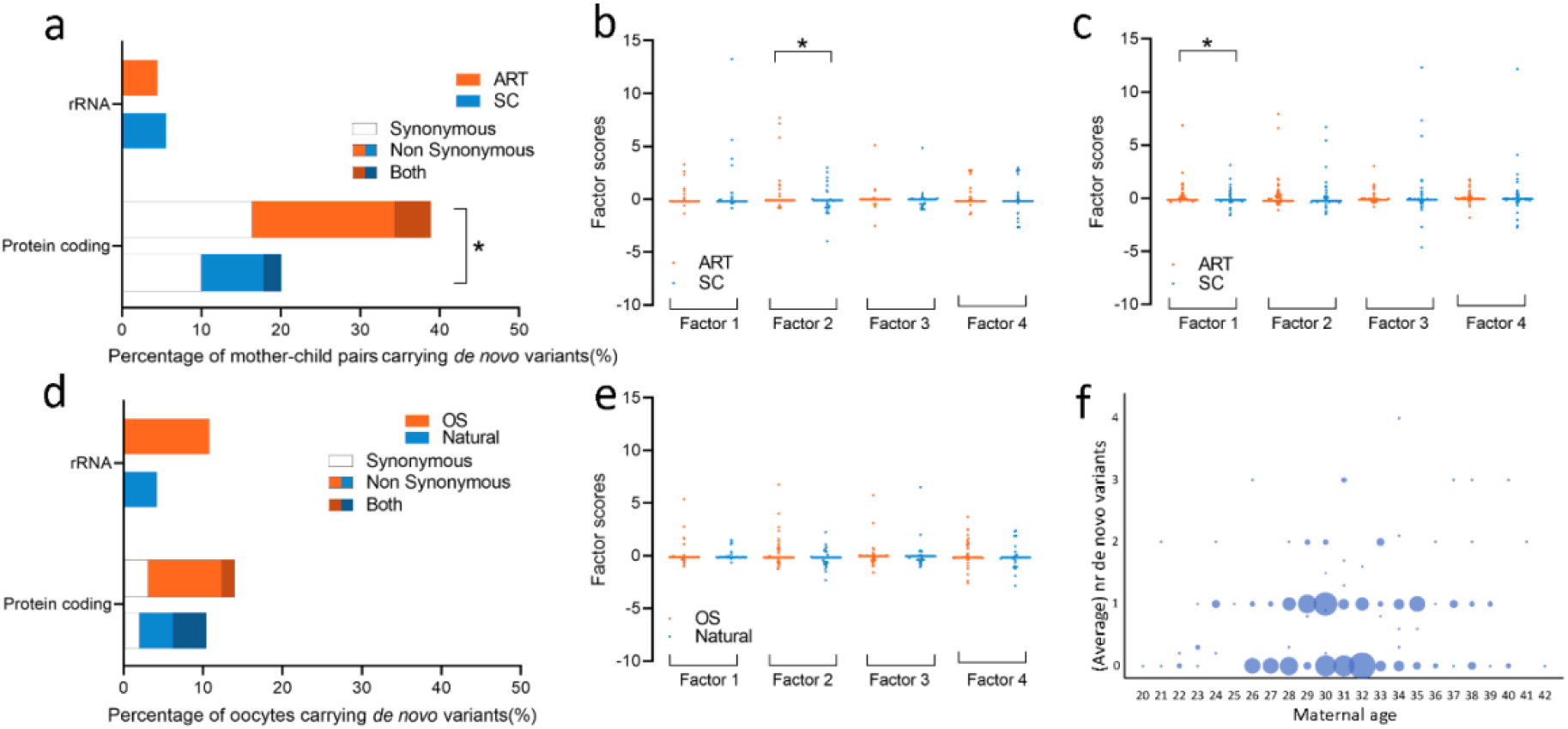
The origin of the differences between ART and SC individuals are mainly due to both maternally transmitted and *de novo* variants and are not caused by ovarian stimulation. **a**. Number of ART and SC mother-child pairs showing *de novo* variants in the protein-coding and rRNA regions. ART children often show more *de novo* protein-coding variants than SC children (*Fisher’s exact test, p=0.01). **b**. Factor scores of the four factors for the transmitted variants in ART and SC children. Factor 2 scores were significantly higher in the ART children (*Mann-Whitney test, p=0.04). **c**. Factor scores of the four factors for the *de novo* variants in ART and SC children. Factor 1 scores were significantly higher in the ART children (*Mann-Whitney test, p=0.03). **d**. Number of oocytes after natural and OS cycles showing *de novo* variants in the protein-coding and rRNA regions. No statistically significant differences were observed (Fisher’s exact tests). **e**. Factor scores of the four factors of the component matrix shown in Figure 1e in natural cycle and OS oocytes. **f**. Poisson Generalized linear model with the total number of *de novo* variants in mother-child pairs and in oocytes plotted against the maternal age at time of conception or oocyte collection. The size of the dots is proportional to the number of samples with the same data point. Maternal ageing correlates significantly with the total number of *de novo* variants in the next generation (p<0.001).

### Maternal ageing rather than ovarian stimulation results in a higher incidence of *de novo* mtDNA variants in the oocyte

Finally, we hypothesized that the ART-associated procedure of ovarian stimulation (OS) may be resulting in the differences in the *de novo* variants that were observed between ART and SC individuals. To test this, we compared the mitochondrial genome of sibling oocytes obtained from either natural menstrual cycles or after OS from twenty-nine young fertile women (Table S1 in supplementary appendix). We sequenced the mtDNA of 113 individual oocytes, 48 from natural and 65 of OS cycles, along with three sources of somatic DNA of the donor. This allowed us to categorize the heteroplasmic variants as either transmitted (present in at least two samples of the same donor) or *de novo* (unique to one oocyte).

Fourteen transmitted variants were identified in the oocytes of seven donors. No trends were observed in changes in heteroplasmic load of the same variant in oocytes obtained after a natural and an OS cycle (Figure S3 in supplementary appendix).

In analogy to the previous analyses, we reduced the complexity of the data by generating the sum of the heteroplasmic loads of the *de novo* variants in each oocyte and was subjecting it to factor analysis (component matrix in Figure S4a in supplementary appendix). To determine whether the *de novo* variants identified in multiple oocytes of the same donor could be considered as independent events in the statistical analysis, we determined the degree of correlation of the score for each factor, for each possible pair of oocytes in each donor. The *de novo* variants in each oocyte of the same donor correlated very poorly with each other, allowing us to conclude that the type and frequency of *de novo* variants is not related to the donor itself and that we could analyse the *de novo* variants in each oocyte as independent events (Figure S4b-d in supplementary appendix). Furthermore, we didn’t find any differences in factor scores between natural and OS cycle oocytes (Figure S4e).

Irrespective of their load, the location of the *de novo* variants as well as the number of oocytes carrying *de novo* protein-coding and rRNA variants, was similar between the natural and OS cycle oocytes (Figure 3d). The sum of the heteroplasmic loads of the *de novo* variants was then analysed using the same matrix as for the children (Figure 1e), in order to test if the *de novo* variants found in the oocytes contributed to the increased factor 2 scores observed in the children. No differences were found in the four factor scores between oocytes from a natural cycle and after OS (Figure 3e). Additionally, we tested the correlation between the total FSH units given at time of treatment and the sum of heteroplasmic loads in the protein-coding regions and the correlations between factor 1 and factor 2 scores in the oocytes and the total amount of FSH units given at the time of treatment, the number of oocytes retrieved in the OS cycle and the maternal age at time of oocyte pick-up. All seven relationships correlated poorly and were not statistically significant (Figure S4f-l).

Finally, we tested using a Poisson Generalized Linear Model if the number of *de novo* variants in mother-child pairs and in the oocytes correlated to the maternal age at the time of conception or oocyte collection. For the oocytes, we used the average number of *de novo* variants per oocyte for each donor. We found that maternal ageing increased the total number of *de novo* variants (p<0.001, B=0.037, 95% Wald CI 0.024-0.05, Figure 3f). Interestingly, the maternal age was significantly higher in the ART group (table I, t-test, p<0.0001), potentially explaining why ART children have a higher incidence of de novo variants than their spontaneously conceived peers.

## DISCUSSION

This study is identifies, for the first time, genetic differences between ART and SC children that are predictive for their birth weight percentile. Also, a striking sex-dependent effect in the ART group was observed, where the presence of protein-coding and rRNA mtDNA variants was predictive for the birth weight percentile of the females but not the males. This sex-dependent effect was previously reported for a sub-cohort of our study, where embryos grown in Vitrolife G3 resulted in a higher birth weight compared to Cook medium, with male embryos being more sensitive to the effect of the culture medium^23^. In another study of ART children, males but not females were found to be significantly heavier at birth when subjected to extended *in vitro* embryo culture^24^. In animal models, the resistance against oxidative stress of bovine male embryos is depending on the culture medium^25^, and mouse female pups, but not male, are born with low birth weight if mitochondrial function is reduced during cleavage stage embryo culture^26^. Taken together, this suggests sex-specific sensitivities to environmental and genetic factors during preimplantation development.

The study of our cohort of mother-child pairs showed that the differences between ART and SC individuals are mainly due increased maternally transmitted protein-coding and rRNA variants, in combination with more *de novo* protein coding variants in the ART individuals. We hypothesized that OS could be responsible for this raise in *de novo* variants, but we found no differences between the natural and OS cycle oocytes. Conversely, maternal ageing positively correlated with the number of *de novo* variants found in the next-generation. This finding is in line with the observations of Zaidi et al.^27^ in human oocytes and work in mouse by Burgstaller et al.^28^. This may explain why ART children have a higher incidence of de novo variants, as their mothers were older than those of SC individuals at the time of conception.

Finally, the exact mechanisms for the association between non-disease causing mtDNA variants and birth weight are yet unclear, but the impact of mtDNA variation on health and disease is well-documented^23^. Studies in mice show that maternally transmitted non-disease causing mtDNA mutations result in low birth weight, premature ageing and decreased fertility^29,30^. In the human, *de novo* somatic mutagenesis is well-associated to ageing and mtDNA variation has been suggested to play a role in female infertility^31^. Inherited disease-causing mtDNA variants are linked to low birth weight, obesity and insulin resistance^8,9,10^ and mitochondrial haplogroups have been associated with a lower bioenergetic fitness^32^, subfertility^33^ and obesity^34^. Further, healthy individuals are known to carry heteroplasmic variants, most of which are not considered to be disease-causing^35^. Nevertheless, variants inducing an amino-acid change or alterations in the transcription machinery can decrease the efficiency of the oxidative phosphorylation and increase the production of reactive oxygen species^36^. Our study has identified a link between the presence of mtDNA heteroplasmic variants in protein-coding regions and rRNA loci and lower birth weight, and an association to being born from subfertile parents after an ART treatment. We propose that these variants may result in a modest but still sufficient enough mitochondrial dysfunction to result in a lower birth weight percentile, providing the first evidence for genetic factors that could explain the differences observed between ART and SC individuals^1,2^. The long-term health consequences of these changes remain to be studied to establish how these findings will impact the clinical practice and patient counselling in the future.

## Data Availability

All data produced in the present study are available upon reasonable request to the authors.

## ACKNOWLEDGEMENTS

The authors would like to express their sincere gratitude towards all staff of the Center for Reproductive Medicine in the UZ Brussel for their engagement in this study, and to thank Hatice Satilmis, Jo Bossuyt and Khadija Rhioui for their support in data collection for this paper. We also thank Iain Johnston and Joanna Poulton for the critical reading and valuable scientific contributions on our work.

## FUNDING

This research was funded by the Research Foundation Flanders (FWO), Willy Gepts Research Foundation of the UZ Brussel and the Methusalem Grant to Prof. Dr. Karen Sermon of the Vrije Universiteit Brussel.

## SUPPLEMENTARY APPENDIX

## Supplementary methods

### Exploratory factor analysis

Exploratory factor analysis is a statistical method used to extract the variability among variables in the form of independent latent variables. We hypothesized that the eight variables generated by adding up the heteroplasmic loads of variants of a given type (all variants in the HV, non-coding, OHR, TAS, protein-coding (synonymous and non-synonymous), rRNA-coding and tRNA-coding regions) could potentially be explained by a lower number of underlying variables, or factors. Hence, we carried out exploratory factor analysis using these eight variables as input. The number of finally chosen factors in the model is based on the eigenvalue. In our case, four factors can represent the majority of variance of the eight variables (see scree plot, panel a), and the relative contribution of each variable to each factor is reflected in the component matrix (panel b). Next, we computed the factor scores per sample, for all four factors. To calculate the factor score for a given sample for a given factor, the sample’s standardized score on each variable is multiplied by the corresponding loadings of the variable for the given factor in the component matrix, and these products are added up. (see panel c for an example). As a result, each sample has scores for the four factors, which reflect its mtDNA variant composition.

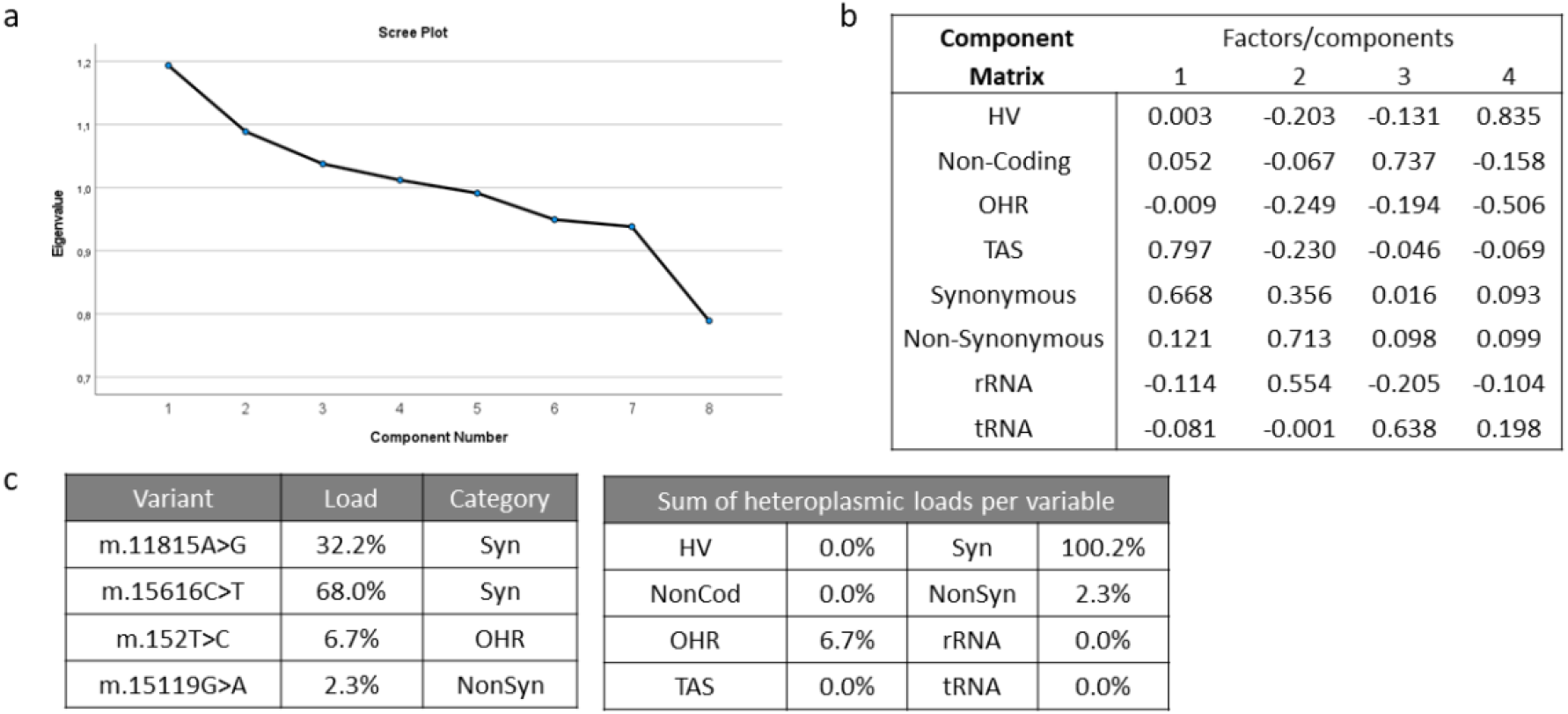

### Binary logistic regression and discriminant analysis

In first instance, we used binary logistic regression by SPSS to investigate the role of the different factors on the probability of being born with a birth weight under the 10th percentile (<P10) and the 25th percentile (<P25). We introduced all confounding factors (the used culture medium, maternal age, maternal BMI >25, gestational hypertension, gestational diabetes, primiparity, and smoking during pregnancy) and the different aspects of the mitochondrial genome (haplogroup, presence of different types of homoplasmic variants, presence of different types of heteroplasmic variants and the values of the factor analysis). Although a number of these factors appeared to have a statistically significant impact, the model generated with the logistic regression was not able to predict the category of birth weight. We therefore considered this logistic regression model not to be sufficiently reliable and choose to work with a discriminant analysis approach which resembles multiple regression analysis. During the building of the model using discriminant analysis in SPSS, we systematically tested the impact of each factor on the accuracy of the model. Overall, factors that proved to be statistically significantly associated to <P10 or <P25, also significantly improved the model. In order to achieve the strongest possible model, we aimed at building a model that would work for both ART and SC individuals. The inclusion of a maximum number of cases resulted in the strongest p-value for the model.

## Supplementary figures and tables

**Figure S1.**
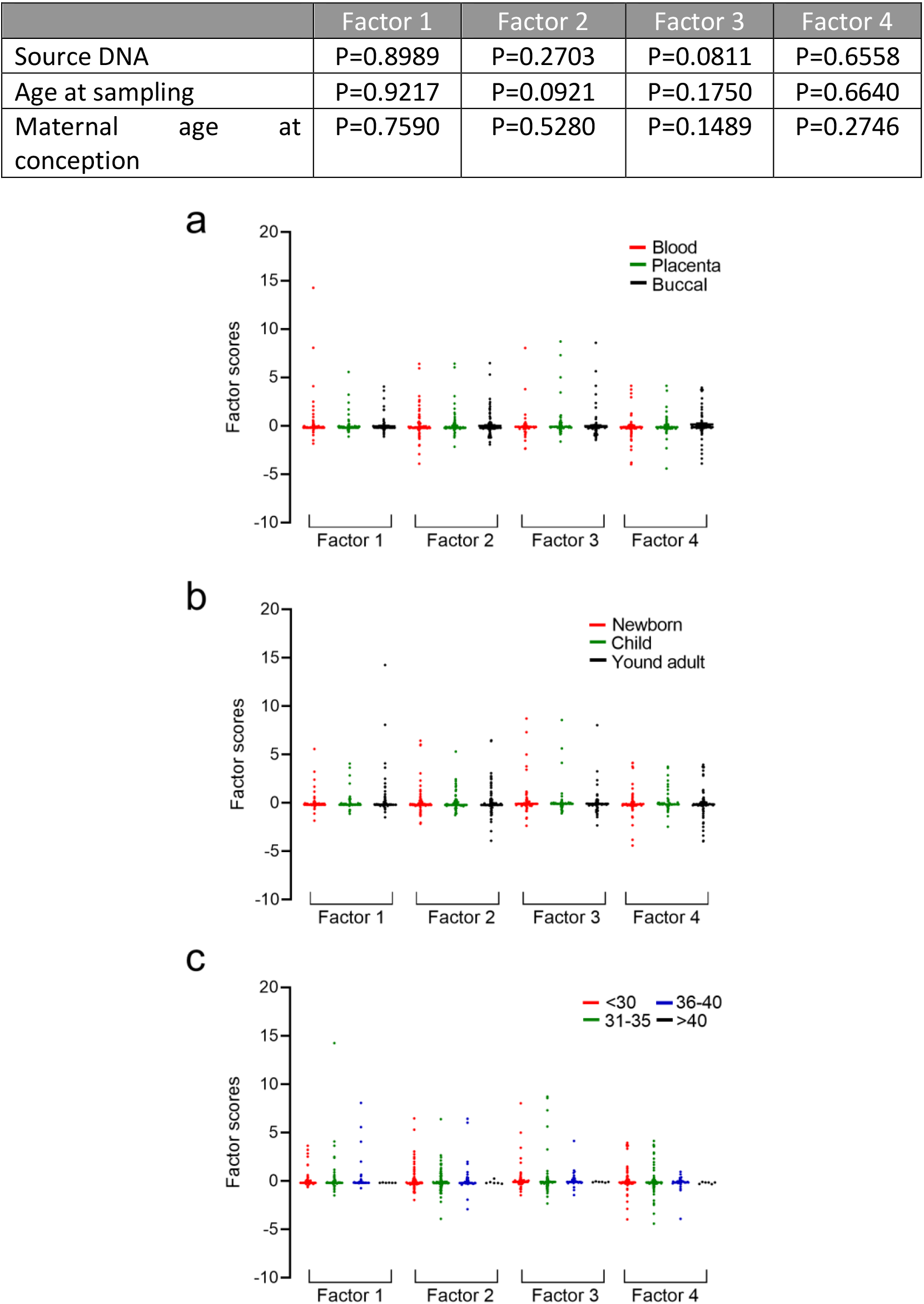
Factors that could potentially influence the presence and load of variants. Factors 1-4 of the factor analysis are plotted according to (**a**) source of DNA (blood: N=168, placenta: N=117, buccal: N=166), (**b**) age at sampling (newborn: N=163, children: N=106, young adults: N=182) and (**c**) maternal age at conception (<30: N=158, 31-35: N=163, 36-40: N=57, >40: N=6). No significant differences were found. Statistics were performed using the Kruskal-Wallis test and were considered significant when p<0.05.

**Figure S2.**
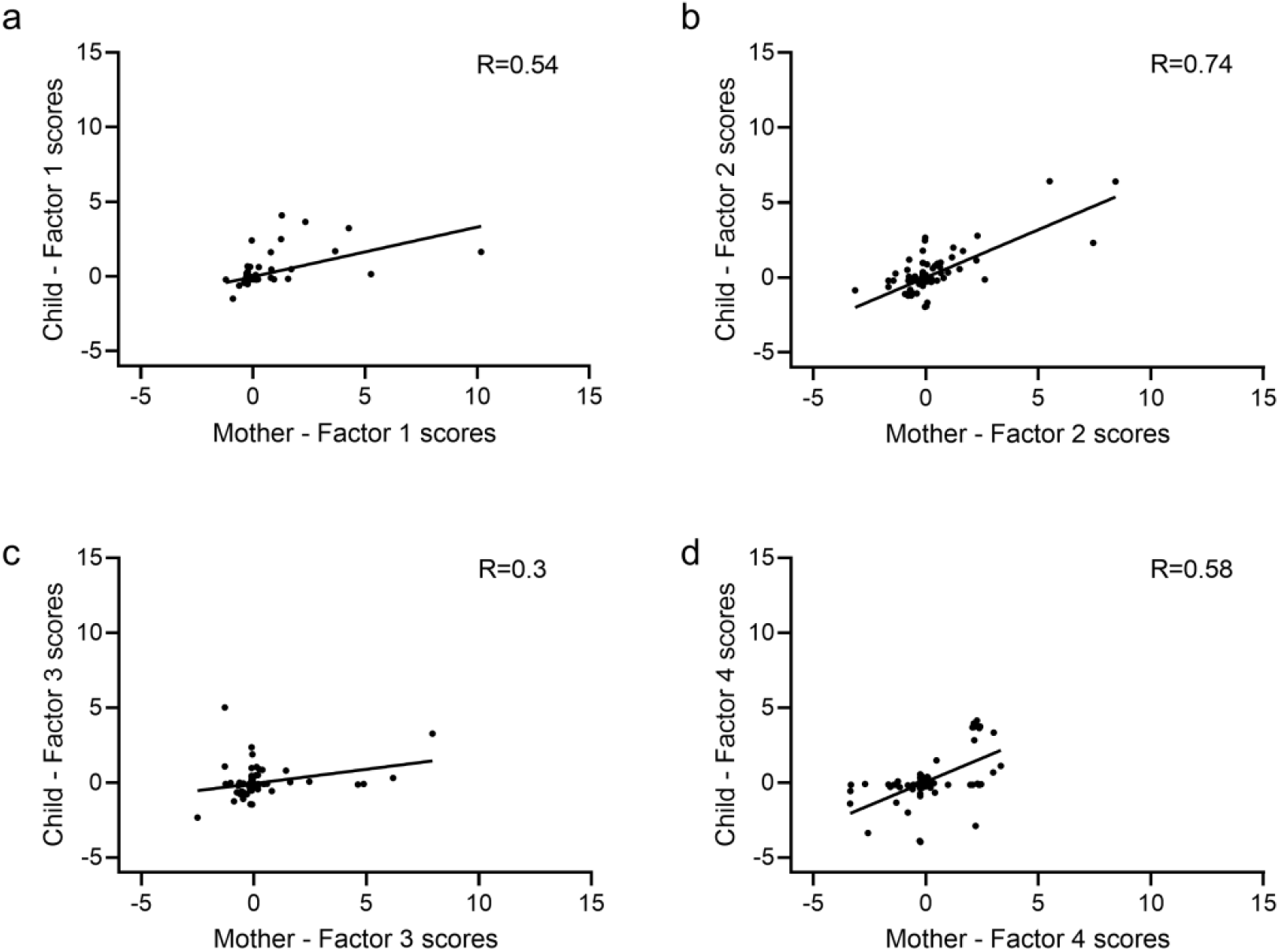
Correlation plots of the factor scores between mother and child. **a-d**. Correlation plot for factors 1, 2, 3 and 4. All correlations were statistically significant (factor 1: p<0.0001, factor 2: p<0.0001, factor 3: p=0.0002 and factor 4: p<0.0001).

**Figure S3.**
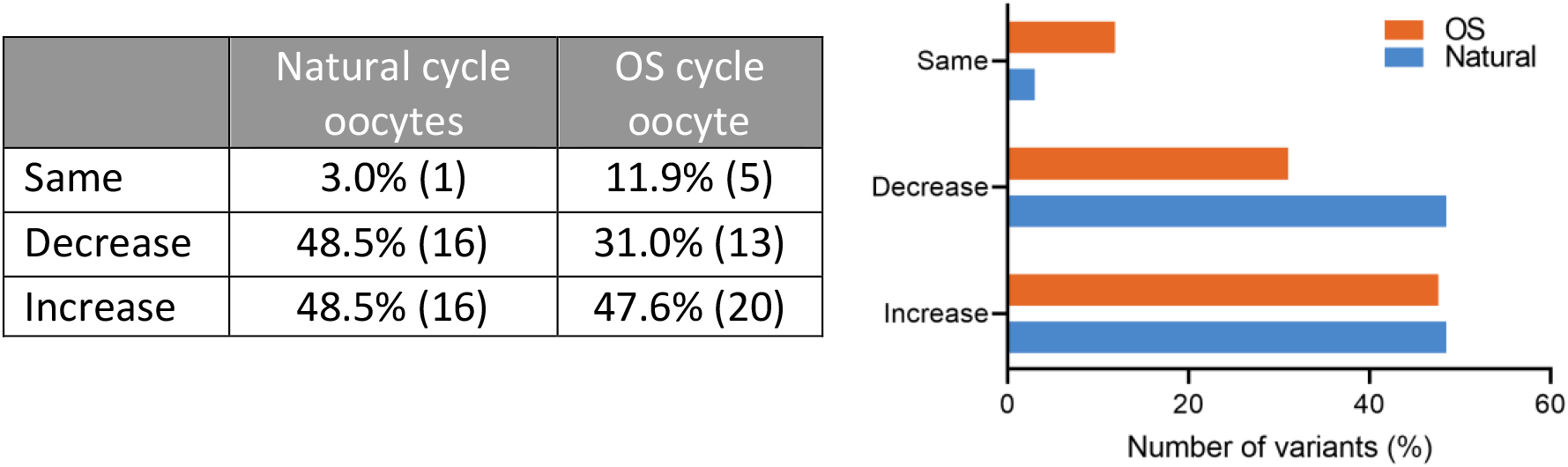
Changes in heteroplasmic load after transmission seen in the oocytes of natural and OS cycles from the same donor. No differences were seen between natural and stimulated oocytes.

**Figure S4.**
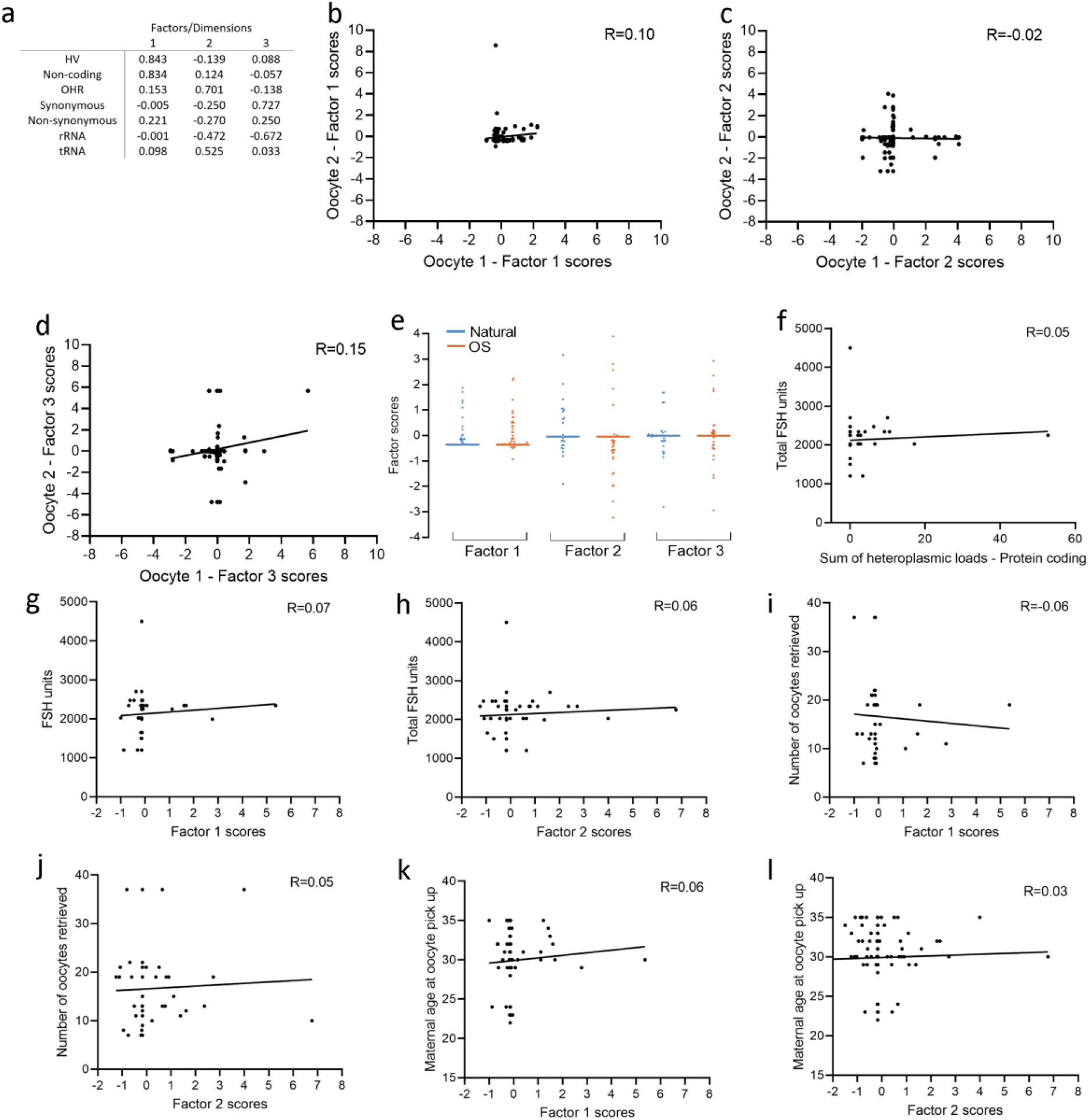
Overview of factor analysis of the oocytes from natural cycles and after OS. **a**. Component matrix generated by the factor analysis using the “sum of the heteroplasmic loads” per oocyte in 7 different categories. **b-d**. Correlation plots for all three factors between oocytes from the same donor for *de novo* variants. All correlations were very low and not significant (Factor 1: p=0.2375, factor 2: p=0.8177 and factor 3: p=0.5110). **e**. Factor scores of the heteroplasmic data of oocytes from natural cycles and after OS. None of the factors were different (Mann-Whitney test, factor 1: p=0.33, factor 2: p=0.07 and factor 3: p=0.57). **f**. Correlation plot of the total amount of FSH units used during treatment and the cumulative heteroplasmic load of the protein-coding variants (p=0.68). **g**. Correlation plot of the total amount of FSH units used during treatment and the factor 1 scores (p=0.58). **h**. Correlation plot of the number of oocytes retrieved at the oocyte pick up and the factor 1 scores (p=0.64). **i**. Correlation plot of the maternal age at the time of oocyte pick up and the factor 1 scores (p=0.54). **j**. Correlation plot of the total amount of FSH units used during treatment and the factor 2 scores (p=0.65). **k**. Correlation plot of the number of oocytes retrieved at the oocyte pick up and the factor 2 scores (p=0.7). **l**. Correlation plot of the maternal age at the time of oocyte pick up and the factor 2 scores (p=0.78). OS: ovarian stimulation, FSH: follicle stimulating hormone.

**Table S1.** Total number of oocytes retrieved in natural and OS cycles per donor. Each donor underwent up to three natural menstrual cycle and one cycle after OS. The oocytes from an OS cycle used in this study were supernumerary to the mature oocytes that were suitable for oocyte donation.

| Donor | Natural cycle oocytes | OS cycle oocytes |
| --- | --- | --- |
| 1 | 3 | 7 |
| 2 | 2 | 2 |
| 3 | 1 | 2 |
| 4 | 1 | 0 |
| 5 | 2 | 6 |
| 6 | 2 | 2 |
| 7 | 3 | 4 |
| 8 | 1 | 0 |
| 9 | 1 | 6 |
| 10 | 1 | 3 |
| 11 | 1 | 3 |
| 12 | 2 | 0 |
| 13 | 3 | 0 |
| 14 | 1 | 2 |
| 15 | 1 | 2 |
| 16 | 3 | 0 |
| 17 | 3 | 7 |
| 18 | 3 | 0 |
| 19 | 3 | 0 |
| 20 | 3 | 4 |
| 21 | 3 | 2 |
| 22 | 3 | 2 |
| 23 | 0 | 0 |
| 24 | 1 | 4 |
| 25 | 1 | 2 |
| 26 | 1 | 2 |
| 27 | 0 | 0 |
| 28 | 0 | 0 |
| 29 | 1 | 3 |

**Table S2.** List of excluded mtDNA positions. Most of the excluded variants were recurrently present across samples. They are present in a homopolymeric stretch and are therefore prone to PCR/sequencing artifacts. The first 250 bp of the amplicons were excluded as well since their coverage was remarkably higher than the rest of the mtDNA sequence, hereby increasing the risk for calling an inaccurate heteroplasmic load.

| Position | Region | Reason |
| --- | --- | --- |
| 303 | MT-HV2; MT-OHR | Recurrent |
| 310-316 | MT-HV2; MT-OHR | Recurrent |
| 513-515 | Non-coding | Recurrent |
| 528-778 | MT-TFH; MT-HSP1; MT-TF; MT-RNR1 | First 250bp of forward primer |
| 955 | MT-RNR1 | Recurrent |
| 1174-1424 | MT-RNR1 | First 250bp of reverse primer |
| 5042-5292 | MT-ND2 | First 250bp of forward primer |
| 5539-5789 | MT-TW; MT-TA; MT-TN; MT-OLR; MT-TC | First 250bp of reverse primer |
| 6286 | MT-CO1 | Recurrent |
| 10935 | MT-ND4 | Recurrent |
| 12148 | MT-TH | Recurrent |
| 13762 | MT-ND5 | Recurrent |
| 15399 | MT-CYB | Recurrent |
| 15407 | MT-CYB | Recurrent |
| 16180-16199 | MT-HV1 | Recurrent |

**Table S3.** Prevalence of haplogroups per mode of conception. Only the haplogroups that were present in more than 10 children were further considered. Statistics were performed using the Fisher’s exact test.

| Haplogroup | ART<br>N=270 | SC<br>N=181 | F test |
| --- | --- | --- | --- |
|  | %(N) |  |  |
| B | 0.4% (1) | 0.6% (1) |  |
| C | 0.4% (1) | 0.0% (0) |  |
| F | 0.4% (1) | 0.6% (1) |  |
| H | 42.6% (115) | 43.6% (79) | P=0.8465 |
| HV | 3.3% (9) | 3.9% (7) | P=0.7988 |
| I | 1.9% (5) | 3.3% (6) | P=0.3612 |
| J | 10.7% (29) | 8.3% (15) | P=0.4224 |
| K | 9.3% (25) | 8.8% (16) | P=1.0000 |
| L | 1.1% (3) | 0.6% (1) |  |
| M | 1.9% (5) | 0.0% (0) |  |
| N | 0.0% (0) | 1.1% (2) |  |
| R | 0.4% (1) | 0.6% (1) |  |
| T | 8.5% (23) | 11.6% (21) | P=0.3316 |
| U | 15.2% (41) | 11.0% (20) | P=0.2611 |
| V | 1.5% (4) | 2.2% (4) |  |
| W | 0.4% (1) | 2.2% (4) |  |
| X | 1.9% (5) | 1.7% (3) |  |
| Y | 0.4% (1) | 0.0% (0) |  |

**Table S4.** Prevalence of subhaplogroups per mode of conception. Statistics were performed using the Fisher’s exact test.

| Subhaplogroups | ART | SC | F test |
| --- | --- | --- | --- |
| H1 | 16.3% (44) | 15.5% (28) | P=0.8958 |
| H10 | 3.0% (8) | 1.1% (2) | P=0.3281 |
| H2 | 1.5% (4) | 3.9% (7) | P=0.1264 |
| H3 | 3.3% (9) | 3.9% (7) | P=0.7988 |
| H5 | 3.0% (8) | 5.5% (10) | P=0.2203 |
| HV0 | 2.2% (6) | 2.2% (4) | P=1.0000 |
| J1 | 7.0% (19) | 4.4% (8) | P=0.3132 |
| J2 | 3.7% (10) | 3.9% (7) | P=1.0000 |
| K1 | 7.8% (21) | 7.7% (14) | P=1.0000 |
| T1 | 2.2% (6) | 4.4% (8) | P=0.2673 |
| T2 | 6.3% (17) | 7.2% (13) | P=0.7047 |
| U4 | 5.2% (14) | 0.6% (1) | P=0.0068 |
| U5 | 7.8% (21) | 7.7% (14) | P=1.0000 |

**Table S5.** Distribution ART and SC individuals with homoplasmic variants outside of the haplogroup. The sum of all the percentages does not add up to 100% because individuals can have multiple homoplasmic variants in the different categories. Synonymous and non-synonymous variants are subcategories of protein-coding variants. Statistics were performed using the Fisher’s exact test.

| Region | ART | SC | F test |
| --- | --- | --- | --- |
| HV | 27.4% (74) | 34.3% (62) | P=0.1427 |
| Non coding | 3.3% (9) | 1.1% (2) | P=0.2124 |
| OHR | 17.4% (47) | 14.9% (27) | P=0.5186 |
| TAS | 1.5% (4) | 0.6% (1) | P=0.6525 |
| Synonymous | 44.4% (120) | 44.8% (81) | P=1.0000 |
| Non synonymous | 27.0% (73) | 34.3% (62) | P=0.1157 |
| rRNA | 9.3% (25) | 5.5% (10) | P=0.1560 |
| tRNA | 7.4% (20) | 5.5% (10) | P=0.5637 |
| Total | 77.4% (209) | 80.1% (145) | P=0.5592 |

**Table S6.** Location of the heteroplasmic variants. Note that the total sum of the absolute numbers of each category does not add up to the total number of variants found in the ART and SC group because the insertions and deletions (ART: n=7 and SC: n=6) in the protein-coding regions are not subcategorized in the synonymous and non-synonymous categories. Statistics were performed using the Fisher’s exact test.

|  | Total | ART | SC | F test |
| --- | --- | --- | --- | --- |
| HV | 19.1% (82) | 17.7% (45) | 21.0% (37) | P=0.4540 |
| Non coding | 4.9% (21) | 4.7% (12) | 5.1% (9) | P=1.0000 |
| OHR | 14.2% (61) | 13.4% (34) | 15.3% (27) | P=0.5767 |
| TAS | 1.2% (5) | 1.2% (3) | 1.1% (2) | P=1.0000 |
| Synonymous | 18.6% (80) | 18.5% (47) | 18.8% (33) | P=1.0000 |
| Non Synonymous | 24.0% (105) | 26.8% (68) | 19.9% (35) | P=0.1085 |
| rRNA | 8.4% (36) | 8.7% (22) | 8.0% (14) | P=0.8607 |
| tRNA | 6.7% (29) | 6.3% (16) | 7.4% (13) | P=0.6982 |

**Table S7.** Type of heteroplasmic variants. Statistics were performed using the Fisher’s exact test.

|  | Total | ART | SC | F test |
| --- | --- | --- | --- | --- |
| Transition | 89.1% (383) | 89.4% (227) | 88.6% (156) | P=0.8754 |
| Transversion | 3.5% (15) | 3.1% (8) | 4.0% (7) | P=0.7903 |
| Insertion | 5.1% (22) | 4.7% (12) | 5.7% (10) | P=0.6627 |
| Deletion | 2.3% (10) | 2.8% (7) | 1.7% (3) | P=0.5367 |

**Table S8.** Mean sum of heteroplasmic loads of the individuals categorized according to their location. Statistics were performed using the Student t test.

|  | ART | SC | Student t test |
| --- | --- | --- | --- |
| HV | 4.5 ± 18.2% | 7.0 ± 23.1% | P=0.2016 |
| NonCod | 0.9 ± 7.6% | 1.1 ± 8.0% | P=0.8068 |
| OHR | 2.1 ± 11.8% | 2.5 ± 11.6% | P=0.7123 |
| TAS | 0.0 ± 0.6% | 0.7 ± 5.8% | P=0.0736 |
| Syn | 2.8 ± 11.5% | 2.2 ± 8.8% | P=0.5798 |
| NonSyn | 2.7 ± 9.7% | 2.1 ± 7.2% | P=0.4910 |
| rRNA | 1.6 ± 9.3% | 1.2 ± 5.5% | P=0.6216 |
| tRNA | 0.9 ± 7.3% | 0.9 ± 4.5% | P=0.9303 |

**Table S9.** (Sub)haplogroup distribution in ART and SC children distributed whether their birth weight was categorized under or above the 25^th^ percentile. Children with haplogroup T were underrepresented in the <P25 category (p=0.04). No significant differences were found. Statistics were performed using the Fischer’s exact test.

| Haplogroups | <P25 | >P25 | P-value |
| --- | --- | --- | --- |
| H | 38.8% (26) | 44.5% (143) | P=0.4186 |
| HV | 6.0% (4) | 3.1% (10) | P=0.2759 |
| I | 4.5% (3) | 1.9% (6) | P=0.1906 |
| J | 14.9% (10) | 9.3% (30) | P=0.1856 |
| K | 9.0% (6) | 9.3% (30) | P=1.0000 |
| T | 3.0% (2) | 11.2% (36) | P=0.0410 |
| U | 14.9% (10) | 11.2% (38) | P=0.5399 |
| Subhaplogroups |  |  |  |
| H1 | 11.9% (8) | 16.8% (54) | P=0.3651 |
| H10 | 3.0% (2) | 2.2% (7) | P=0.6570 |
| H2 | 1.5% (1) | 3.1% (10) | P=0.6978 |
| H3 | 4.5% (3) | 3.1% (10) | P=0.4769 |
| H5 | 3.0% (2) | 4.0% (13) | P=1.0000 |
| HV0 | 4.5% (3) | 1.6% (5) | P=0.1443 |
| J1 | 9.0% (6) | 5.6% (18) | P=0.2757 |
| J2 | 6.0% (4) | 3.7% (12) | P=0.4947 |
| K1 | 7.5% (5) | 7.8% (25) | P=1.0000 |
| T1 | 0.0% (0) | 4.0% (13) | P=0.1367 |
| T2 | 3.0% (2) | 7.2% (23) | P=0.2786 |
| U4 | 4.5% (3) | 2.5% (8) | P=0.4120 |
| U5 | 4.5% (3) | 8.1% (26) | P=0.4440 |

**Table S10.**
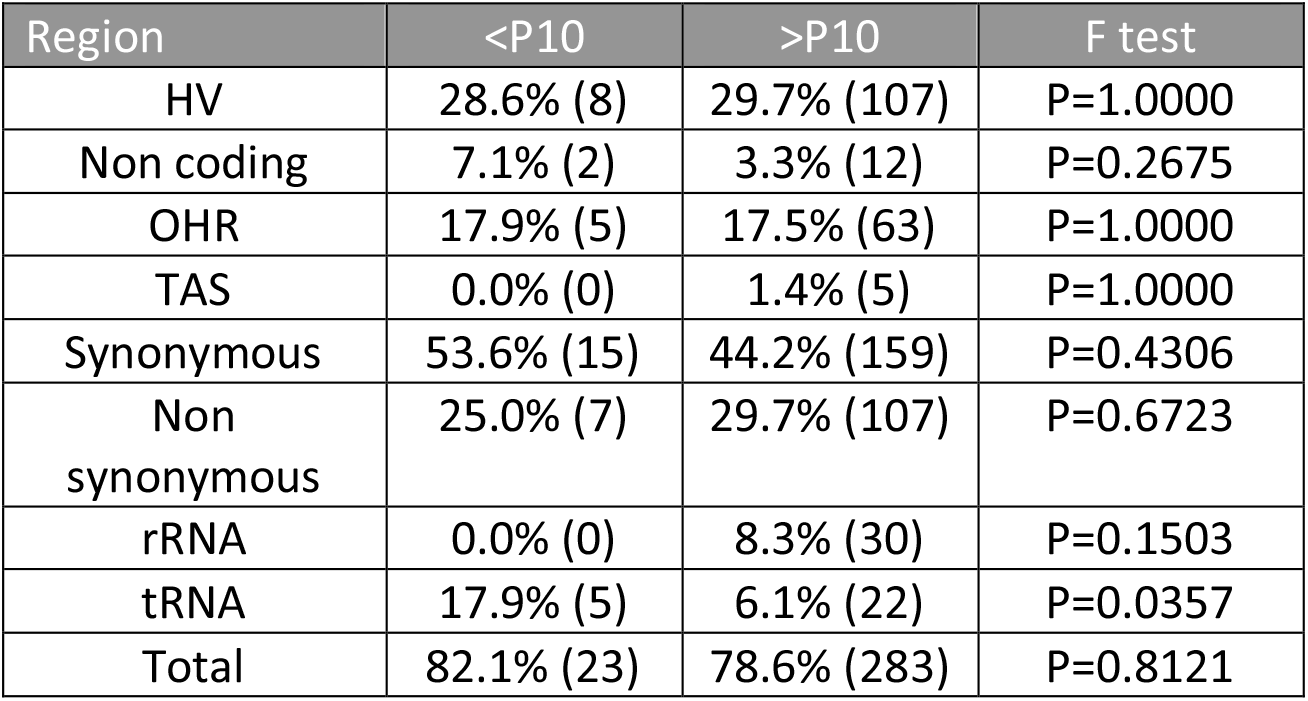
Distribution ART and SC individuals with homoplasmic variants outside of the haplogroup distributed whether their birth weight was categorized under or above the 10^th^ percentile. The percentages do not add up to 100% because individuals can have multiple homoplasmic variants in different categories. Statistics were performed using the Fisher’s exact test.

**Table S11.**
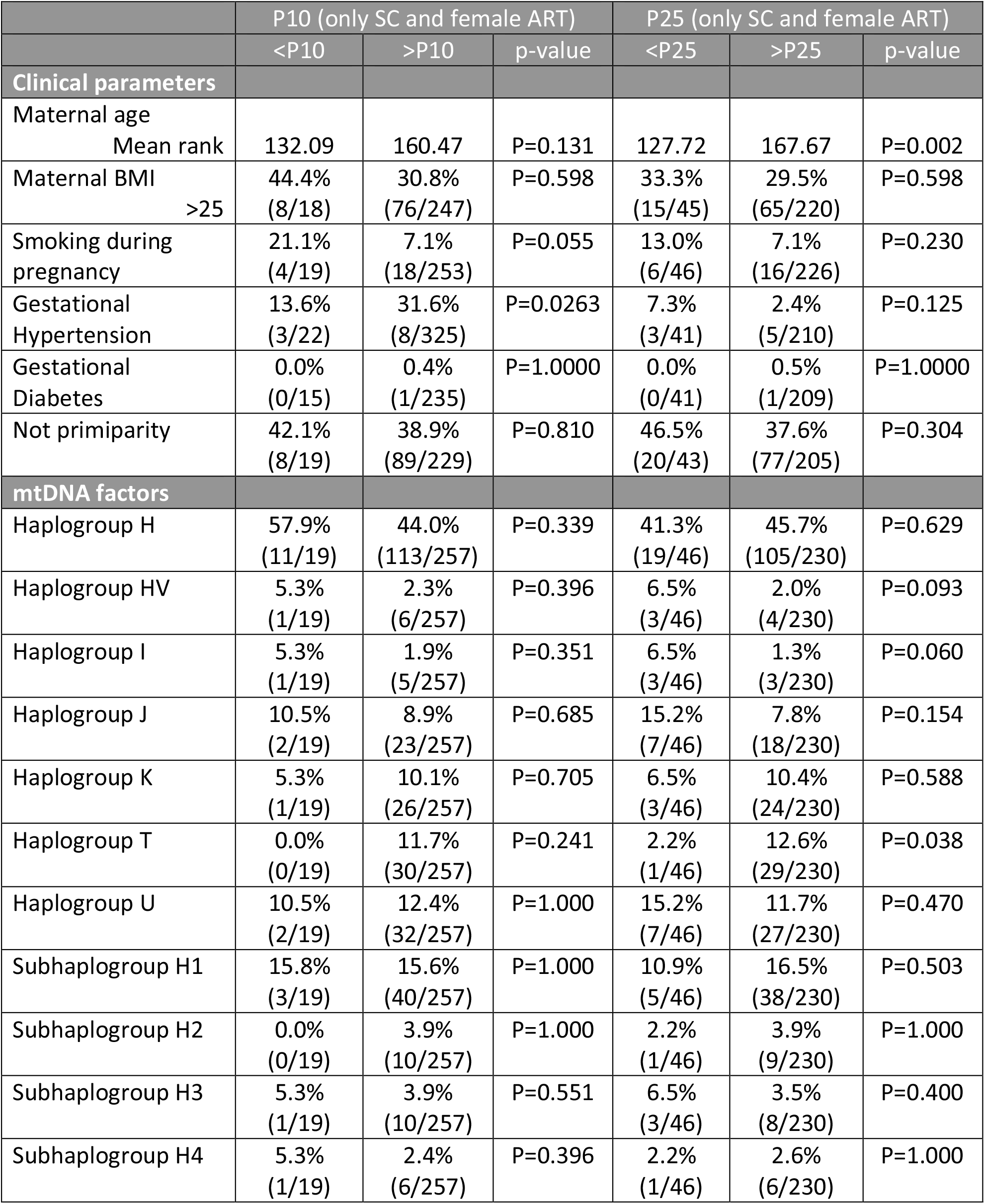
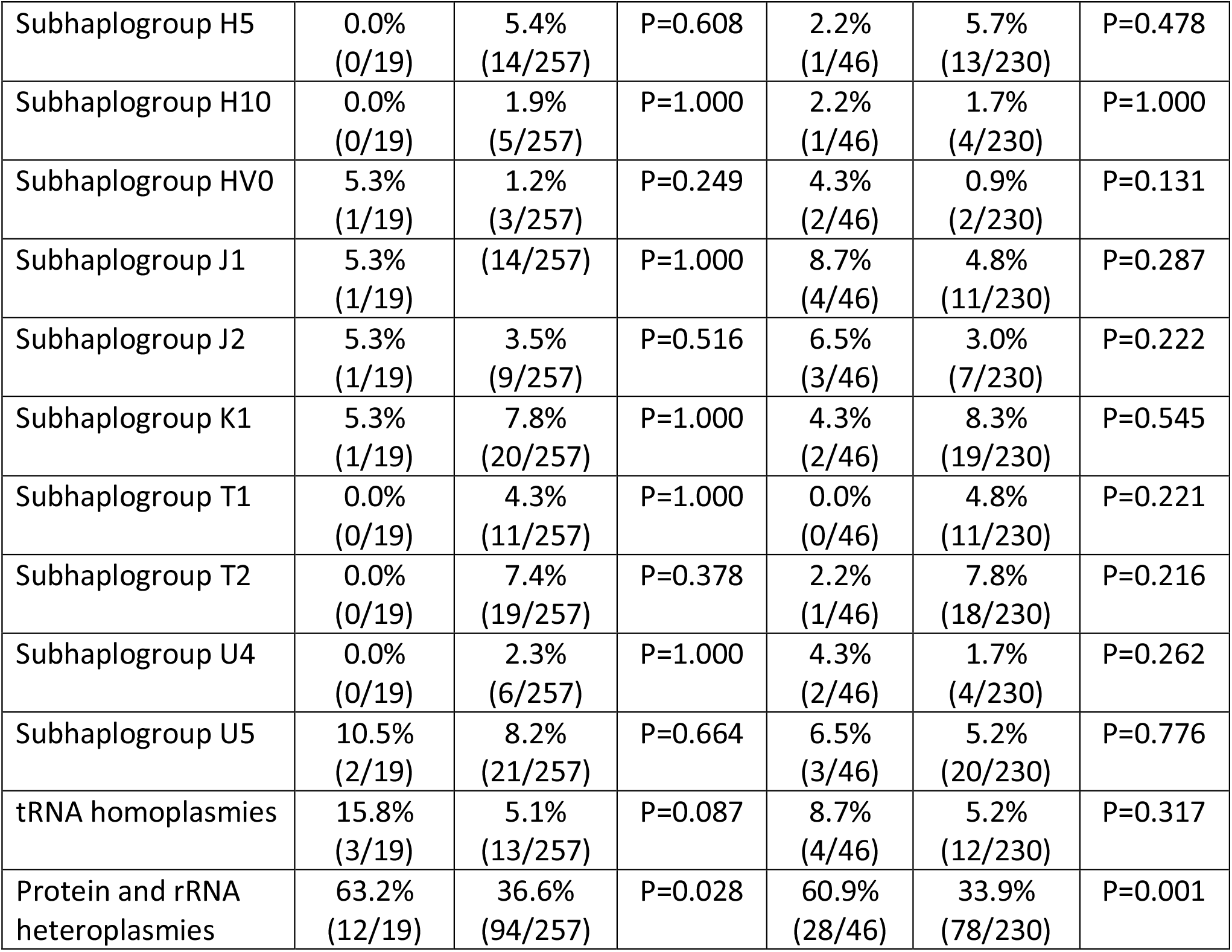
List of possible confounding factors affecting birth weight. Statistical tests were performed with Chi-square test, except for maternal age which was done with a Mann-Whitney test. Parameters with a p-value <0.2 were included in further statistical models, except for subhaplogroup HV0 in the P25 as this would cause a bias with haplogroup HV as a parameter.

## Notes

### Competing Interest Statement

The authors have declared no competing interest.

### Funding Statement

This study was funded by the Research Foundation Flanders (FWO), Willy Gepts Research Foundation of the UZ Brussel and the Methusalem Grant to Prof. Dr. Karen Sermon of the Vrije Universiteit Brussel.

### Author Declarations

The local ethical committee of the University Hospital UZ Brussel/Vrije Universiteit Brussel and the ethical review board of the Maastricht University Medical Centre gave ethical approval for this work

